# Functional activation changes at ultra-high field related to upper and lower limb impairments in multiple sclerosis

**DOI:** 10.1101/2020.08.13.20174664

**Authors:** Myrte Strik, Camille J. Shanahan, Anneke van der Walt, Frederique M. C. Boonstra, Rebecca Glarin, Mary P. Galea, Trevor J. Kilpatrick, Jeroen J.G. Geurts, Jon O. Cleary, Menno M. Schoonheim, Scott C. Kolbe

## Abstract

Upper and lower limb impairments are common in people with multiple sclerosis (pwMS), yet difficult to clinically identify in early stages of disease progression. Tasks involving complex motor control can potentially reveal more subtle deficits in early stages, and can be performed during functional MRI acquisition, to investigate underlying neural mechanisms, providing markers for early motor progression. We investigated brain activation during visually-guided force-matching of hand or foot in 28 minimally disabled pwMS and 17 healthy controls (HC) using ultra-high field 7-Tesla fMRI, allowing us to visualise sensorimotor network activity in high detail. Task activations and performance (tracking lag and error) were compared between groups, and correlations were performed. PwMS showed delayed (+124 s, *p*=0.002) and more erroneous (+0.15 N, *p*=0.001) lower limb tracking, together with higher primary motor and premotor cortex activation, and lower cerebellar activation compared to HC. No differences were seen in upper limb performance or activation. Functional activation levels of cerebellar, visual and motor areas correlated with task performance. These results demonstrate that ultra-high field fMRI during complex hand and foot tracking can identify subtle impairments in movement and brain activity, and differentiates upper and lower limb impairments in minimally disabled pwMS.

## Introduction

Multiple sclerosis (MS) is a progressive disease of the central nervous system characterized by neuroaxonal inflammation, demyelination and degeneration. Motor impairments are common and disabling, and include tremor, muscle weakness, loss of fine motor control, ataxia and loss of mobility.(Coghe et al., 2019) Walking difficulties are a typical hallmark of MS and occur in most people with MS (pwMS) (80%) within 10 to 15 years(Souza et al., 2010), and upper limb impairments also eventually occur in up to 80% as well,(Johansson et al., 2007) yet are less well recognised and studied. Also, the degree of impairment of upper and lower limbs correlates only moderately,(Coghe et al., 2019) suggesting at least partially divergent mechanisms of progression. Elucidating such mechanisms is important for formulating personalised treatment strategies, especially early in the disease when clinical signs of disability are minimal yet efficacious treatment has the best chance of avoiding significant neurological decline.

Functional MRI (fMRI) provides a useful means of studying changes in brain activity and putative neural compensations that contribute to the rate of progression of motor impairments. Task-based fMRI reveals regional brain activation indirectly via local changes in blood flow in the cortex driven by neural activity. Such studies have shown that changes in activation can predate changes in sensorimotor impairments and could provide early indicators of subsequent neurological decline.(Filippi et al., 2004) Multiple studies in MS have reported altered patterns of brain activation with increased activation seen in regions typically devoted to simple(Filippi et al., 2004; Mezzapesa, Rocca, Rodegher, Comi, & Filippi, 2008; Pantano et al., 2002; Rocca et al., 2005, 2003) and more complex motor tasks.(Filippi et al., 2004; Rocca et al., 2005) However, a key shortcoming of the extant literature lies in the stronger focus on hand function(Mezzapesa et al., 2008; Pantano et al., 2002; Rocca et al., 2005, 2003) which can easily be performed during scanning with simple tasks involving flexion and extension of the hand or fingers(Mezzapesa et al., 2008; Pantano et al., 2002; Rocca et al., 2005, 2003). Lower limb studies are more difficult to perform and rare, mostly including clinically isolated syndrome (Filippi et al., 2004) or primary progressive MS(Ciccarelli et al., 2006). As such, there is little known about the neural changes accompanying tasks involving complex motor control that provide better models of day to day motor functions.

This study aimed to investigate changes in complex hand and foot motor control in pwMS. The motor task involved a controlled visually-guided contraction of the ankle dorsiflexors (tibialis anterior) or finger/thumb flexors muscles,(Mayhew, Porcaro, Tecchio, & Bagshaw, 2017; Shanahan, Hodges, Wrigley, Bennell, & Farrell, 2015) chosen to activate a broad network involved in integrating complex proprioceptive and visual inputs to accurate track a moving target. In addition, we used ultra-high field 7 Tesla MRI to perform simultaneous multi-slice fMRI at near anatomical MRI resolution (1.24mm isotropic) with high temporal resolution (1.7 sec) to detect subtle and focal activation changes that are undetectable by clinical field strength imaging systems.

## Methods

### Participants

Twenty-eight participants with relapsing-remitting MS (RRMS) (mean age = 41.9 ± 10.0 years; 23 women) were recruited from the Royal Melbourne Hospital, Melbourne, Australia. At time of recruitment, all patients were diagnosed with clinically definite MS according to the revised 2010 McDonald criteria,(Polman et al., 2011) and did not experience any relapses during the 6 months prior. Clinical disability was assessed using the Expanded Disability Status Scale (EDSS) and we included only pwMS with no to minimal clinical disability, i.e. EDSS < 4, and Kurtzke Functional Systems Scale (FSS) for pyramidal and cerebellar function ≤ 2. Exclusion criteria were: any neurological condition other than MS, orthopaedic conditions causing disability of the lower limbs (including painful osteoarthritis), and coexisting cardiovascular disease. Seventeen healthy controls (HC) (mean age = 39.3 ± 7.3 years; 10 women) with no reported history of neurological disorders were recruited for comparisons. Approval was obtained from the Melbourne Health Human Research Ethics Committee and all participants provided a voluntary written consent prior to participation.

### MRI acquisition

Image acquisition was conducted using a whole body Magnetom 7T MRI system (Siemens Healthcare, Erlangen, Germany) with a single-channel transmit and 32-channel receive head coil (Nova Medical, Wilmington MA, USA). Two runs of functional MRI (fMRI) were acquired (upper and lower limb motor tasks performed in separate runs) with the following parameters: repetition time (TR)=1700 ms, echo time (TE)=34.4 ms, flip angle (FA)=65 degrees, multiband slice acceleration factor=6, Fat suppression, GRAPPA phase acceleration factor=2, 120 slices, 1.24 mm isotropic resolution, 165 volumes, image matrix = 168×168). In addition, a 3 dimensional T1-weighted structural image (MP2RAGE: TR=4900 ms, TE=2.89 ms, inversion time (TI)=700/2700 ms, FA=5/6 degrees, 192 slices, GRAPPA phase acceleration factor = 4, phase encoding direction = AP, voxel size = 0.9 mm isotropic, image matrix 256×256) was used for brain volumetric measurements. A fluid-attenuated inversion recovery (FLAIR) scan was acquired for lesion identification (TR=10000 ms, TE=96 ms, TI=2600 ms, flip angle=145 degrees, 45 axial slices, GRAPPA=3, voxel size = 1.2×1.2×3.0 mm, image matrix 192×192).

### Anatomical MRI processing

Anatomical MRI processing involved lesion detection and brain segmentation. White matter (WM) lesions were detected and marked on the MP2RAGE image using a semi-automated thresholding technique in MRIcron (https://www.nitrc.org/projects/mricron) with the FLAIR image used as a reference to avoid inclusion of CSF. Next, lesion maps were used to lesion fill the MP2RAGE images using SLF software (http://atc.udg.edu/nic/slfToolbox/index.html) in Statistical Parametric Mapping (SPM, version 8), which were subsequently used as input for brain segmentation analyses using FreeSurfer version 6.0-patch (https://surfer.nmr.mgh.harvard.edu). From FreeSurfer analysis, the total white matter, cortical grey matter, deep grey matter and ventricular volumes were used for statistical analysis and normalised by total intracranial volume. Spinal cord cross-sectional area at the level of the superior margin of the odontoid peg was marked manually and measured on MP2RAGE images using Horos (v4.0, www.horosproject.org) (Supplementary Figure 1).

### Pre-MRI testing

Immediately prior to the MRI scan session, the upper and lower limb tasks were explained and practiced in a room outside the 7T scanner using a similar experimental setup as in the scanner. During this practice session, the maximum voluntary contraction (MVC) was performed to tailor contraction intensity to each participant’s strength. The participants were asked to squeeze a sphygmomanometer cuff, used to measure force production, with their fingers or by isometric dorsiflexion against a constraint of their foot as strongly as possible for several seconds. The task was performed at a maximum of 5% of MVC as it was found to activate the movement related network activation with minimal head motion.(Shanahan et al., 2015)

### Force matching task participant and apparatus setup

Participants performed a visually guided force-matching task adapted from a previous study and using same apparatus.(Shanahan et al., 2015) Setup of the participant in the scanner for upper limb task was that the cuff was held in the right hand between the thumb and fingers with the hand resting on the chest for support and to limit head movement. The lower limb task setup involved the following: the participant’s right thigh was supported by the scanner bed, lower leg supported by a pillow and the plantar surface of foot resting against the footplate of a MRI compatible rig, all to restrict movement around the ankle with minimal head motion. A sphygmomanometer cuff was positioned over the dorsum of the right foot as close as possible to the metatarsal heads without covering the toes (Figure 1). Between the upper and lower limb fMRI runs, an experimenter entered the MRI room and removed the cuff from the participant’s right hand and positioned it over the foot.

**Figure 1.**
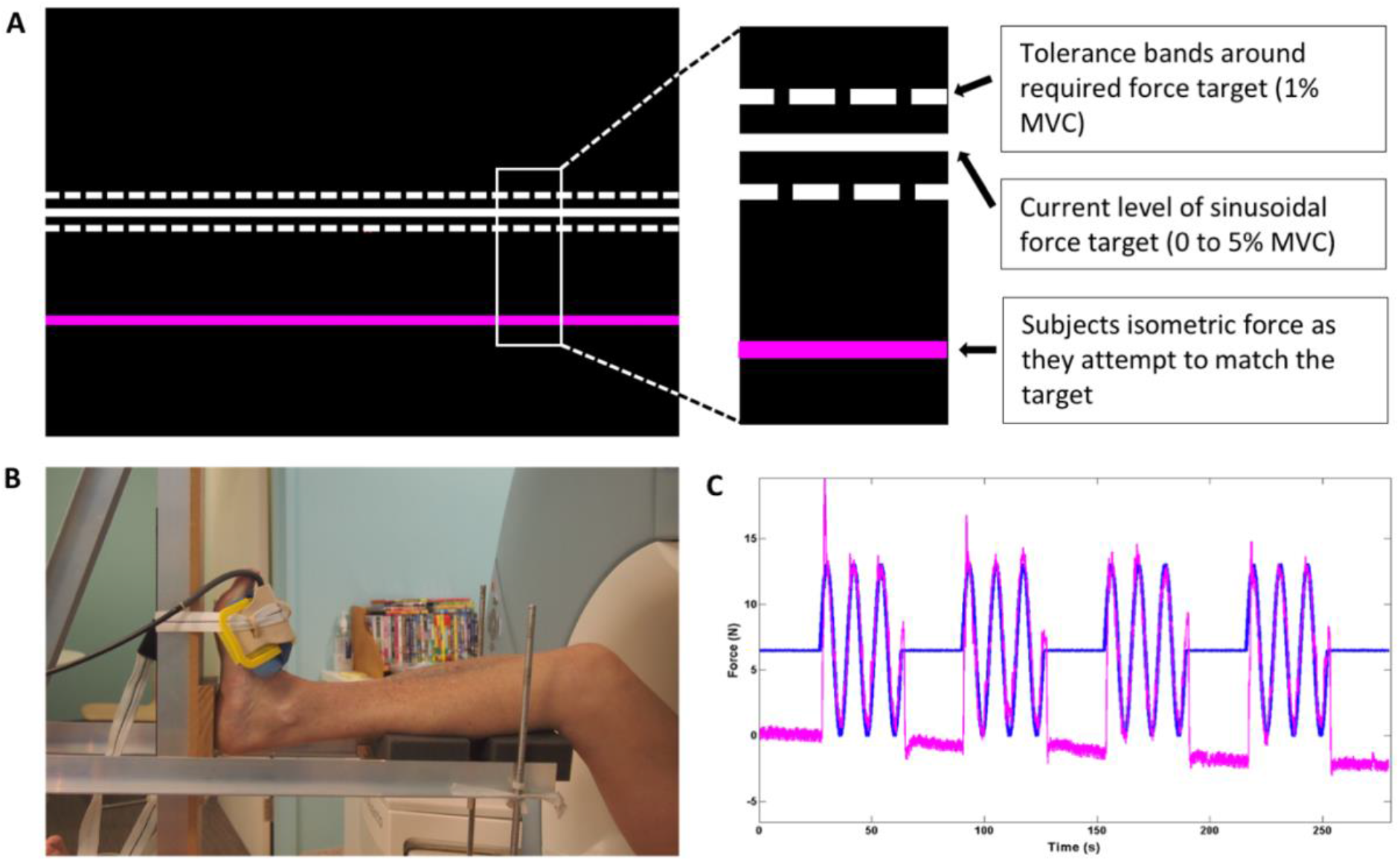
Force-matching task during functional MRI acquisition. **A)** The task presented to the participants. The white line slowly moved up and down during a task block and the participants were asked to follow the white line as accurately as possible with the pink by squeezing and releasing their fingers or pulling their foot up and down to make the pink line hit the white line. **B)** This image shows the MR compatible rig used to stabilise the foot and lower leg during the ankle motor task, i.e. ankle dorsiflexion to match a force indicator displayed to the participant. The sphygmomanometer cuff is positioned over the dorsum of foot close as possible to metatarsal heads without covering toes**. C)** The blue line reflects the force in 4 contraction blocks interleaved with 5 periods of rest. The pink line is an example of the force produced by the participant to match the force indicator displayed on the black screen (white line).

A flowchart showing connectivity of the various apparatus used in the experiment is shown in Supplementary Figure 2. The sphygmomanometer cuff was connected to an air-tight plastic tube and linked through the wave guide to the pressure transducer unit. The pressure transducer was fitted with a manual pressure regulator that was used to fill the apparatus with a reasonable air pressure to allow sensitive pressure change measurements (~40 psi). The transducer was connected to the analogue to digital converter (ADC) via a BNC connected cable. An analogue voltage waveform generator was also connected to the ADC to generate the stimulus waveforms to be tracked. The ADC was connected to the presentation PC via a USB connection. Both target force and the force produced by the participant were displayed (Figure 1) at the rear of the scanner bore using an LCD projector and viewed through a head-coil mounted mirror.

### Visually guided force-matching motor task

The task involved a controlled, low force contraction of either the right ankle dorsiflexors (tibialis anterior) or the right finger/thumb flexor (hand) muscles in independent fMRI runs (Figure 1). Task design involved 4 (45 sec) contraction blocks interleaved with 5 (27 sec) periods of rest. During the contraction block, the force target moved up and down in a simple harmonic motion with a frequency of 0.125 Hz indicating a target contraction intensity from 0 to 5% of MVC. Participants were instructed to match the sinusoidal target force as accurately as possible. The onset of each contraction block was initiated by an MRI trigger to ensure accurate timing between the experiment and MRI data collection.

### Upper and lower limb behavioural task parameters

Two behavioural task parameters (lag and error) were computed from the force data acquired during MRI testing using a custom MATLAB script. Lag was defined as the delay between the task cue and the response produced by the participants. Lag was calculated as the maximum crosscorrelation between the cue and response time series, expressed in milliseconds (ms). The error was defined as the root mean square difference between the cue and the lag-corrected response time series, expressed in Newtons (N).

### Functional MRI pre-processing

Due to the cortical specificity and detailed activation patterns evident at 7T, we chose to create a study-specific template script using Advanced Neuroimaging Tool (ANTS, v.2.3.1, http://stnava.github.io/ANTs/) (Figure 2). The template creation involves 4 iterative coregistrations of an example fMRI volume from each participant. Each co-registration iteration involved rigid body (n=20), affine (n=50) and deformable non-linear (n=20) co-registration calculations followed by averaging of the resultant warped images to create an updated target template. The final template was linearly registered to MNI-152 space for use in fMRI results overlays and coordinate reporting.

**Figure 2.**
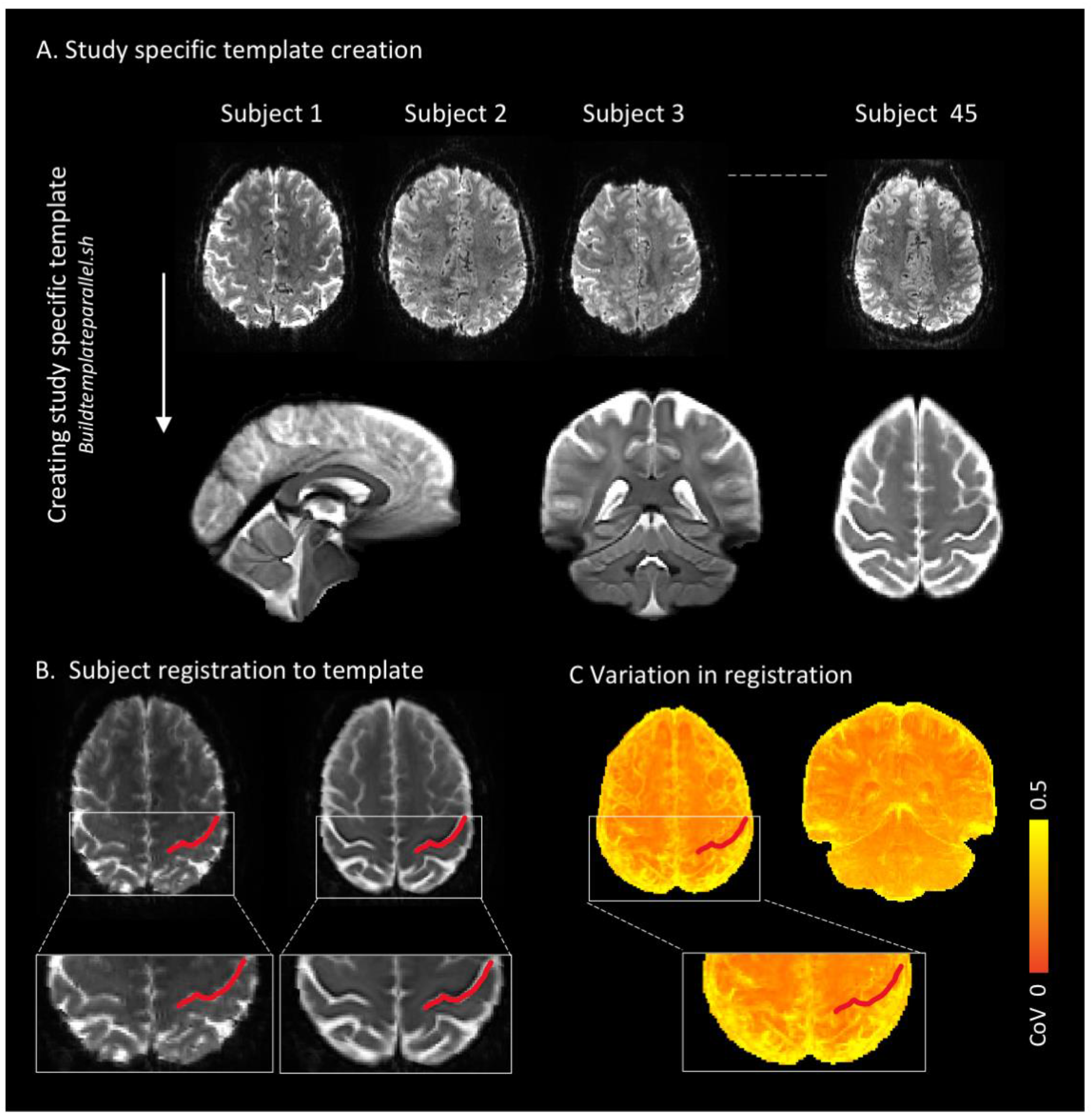
Study-specific template creation and fMRI processing steps. **A)** A study-specific template was created using Advanced Neuroimaging Tools (v.2.3.1, http://stnava.github.io/ANTs/) buildtemplateparallel script and involved 4 iterative coregistrations of an example fMRI volume from each participant (45 participants in total). **B)** An example of a participant to template registration with the red line indicating the central sulcus. **C)** The accuracy of template registrations of all subjects is visualised here. The coefficient of variations (CoV) of the fourth iteration was calculated with yellow reflecting high variability between participants, and towards orange more accurate overlap in registrations.

fMRI data were pre-processed with the following procedures using FEAT (FSL, FMRIB, Oxford, UK, v.6.0.3): rigid body head motion correction (FMRIB Linear Registration Tool), high pass filtering (cut off 100 s) and nonlinear spatial smoothing (SUSAN, extent threshold 2.5 mm) (Figure 2). Volumes affected by excessive motion (mean relative displacement from previous volume > 0.5 mm) were censured from subsequent analyses.

### Functional MRI statistical analyses

We performed a three-level analysis (run, subject, group) using FEAT. Three group-level analyses were performed. Firstly, the main effect of the task was compared between HC and pwMS for upper and lower limbs separately. Where significant clusters were detected, post-hoc correlations were performed to investigate the relationship between the max z-stat values extracted from the clusters and task performance or clinical disability using linear regression and partial rank correlation respectively. Secondly, differences in activation associated specifically with upper or lower limb task performance were investigated. For each participant, subject level analyses were performed on unthresholded run-level activation maps to calculate activation maps specific to either upper or lower limb movements using the following contrasts: upper > lower and lower > upper. Higher level analyses were subsequently performed to calculate significant mean activation maps for upper and lower limbs specifically, and to compare these activation patterns between groups.

Lastly, correlations between mean activation patterns during both tasks and the task parameters (lag and force error) in pwMS were calculated with higher level FEAT analyses with run-level activation maps as input. A mask was used for all correlation analyses to ensure that only brain regions found to be significantly active during the upper or lower limb tasks were included.

All group level statistical analyses employed FLAME 1 mixed effects analysis, and significant voxels were identified using family-wise error correction with a threshold of z-stat > 2.3 and cluster wise significance of p < 0.05.

### Statistical Analyses

Statistical analyses were performed using SPSS (version 26; IBM Corp., Armonk, NY, USA). Demographics, brain volumetrics and task parameters were compared between HC and pwMS with independent sample t-tests, or Mann-Whitney U tests when violations of the assumption of normality occurred (based on Kolmogorov-Smirnov testing and histogram inspection). Linear regressions were performed to investigate relations between the lag and force error of the upper limb and lower limb and between upper and lower limb task performance. The unstandardized residuals were tested for normality using the Kolmogorov-Smirnov test. If non-normally distributed, partial rank correlations were performed. For all correlation analyses, age and sex were included as covariates. P-values were corrected using False Discovery Rate (FDR) and were considered statistically significant < 0.05.

## Results

### Demographics and clinical characteristics

The demographics, clinical measures and brain and lesion volumes are presented in Table 1. PwMS were all diagnosed with RRMS and had a mean disease duration of 6.4 years (SD=3.9), median EDSS of 1.5 (interquartile range (IQR)=1, 1.5). No differences between groups were observed for age, sex or hand dominance. Compared to HC, pwMS displayed significantly reduced white matter volume (*p*=0.002), cortical grey matter volume (*p*=0.014) and deep grey matter volume (*p*=0.005). No differences were observed for ventricular volume and spinal cord cross-sectional area.

**Table 1.**
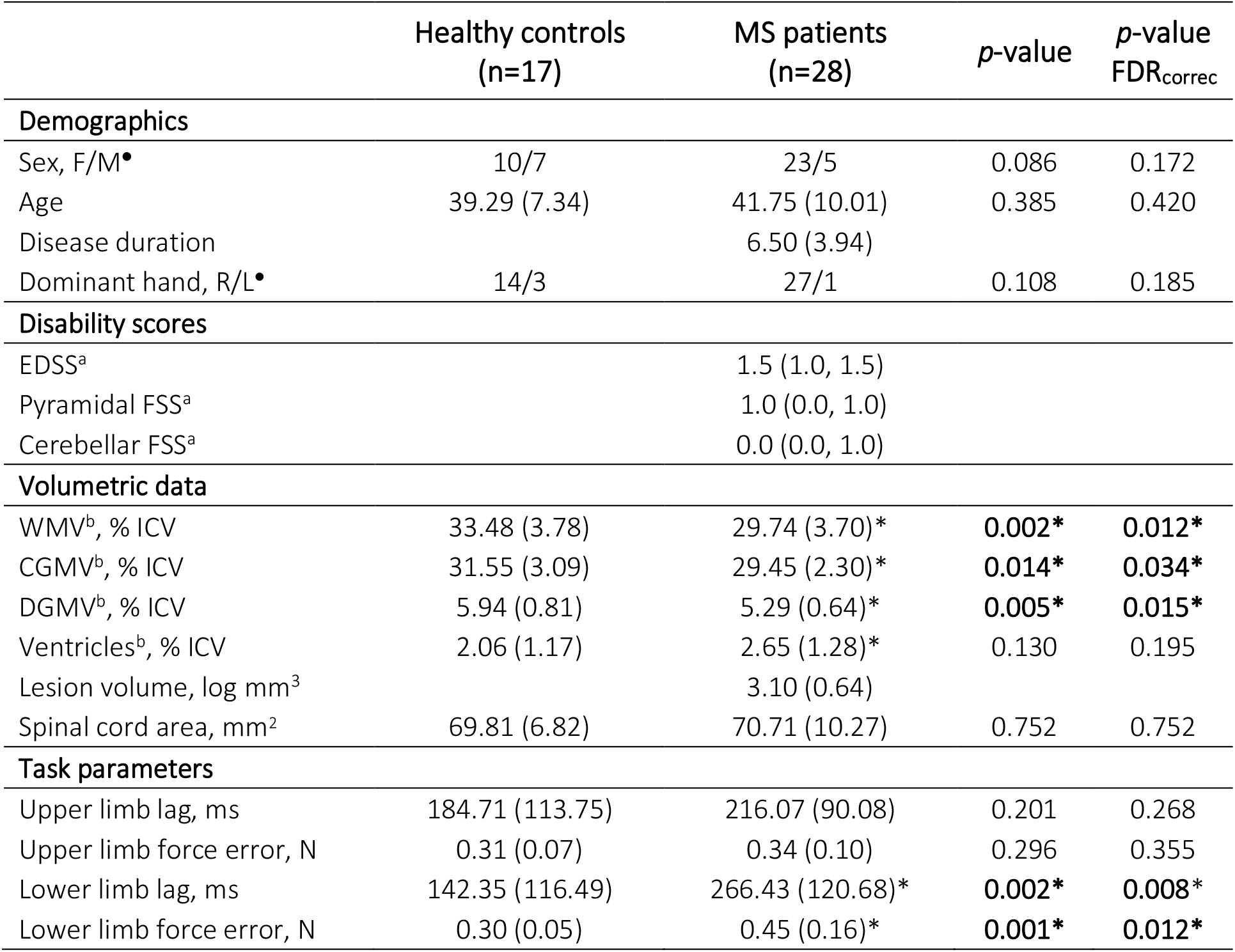
Demographic, clinical, MRI and task performance characteristics. All values represent means and standard deviations, unless denoted otherwise. MS = multiple sclerosis; F = females; M = males; R = right; L = left; EDSS = Expanded Disability Status Scale; FSS = Functional System Score; WMV = white matter volume; ICV = intracranial volume; CGMV = cortical grey matter volume; DGMV = deep grey matter volume. All variables were tested using independent samples t-test and values represent means and standard deviations unless denotes otherwise. ^a^Median and interquartile range. ^b^Brain volumes were normalised for intracranial volume. •Chi-Square test. ^*^Significant difference between people with multiple sclerosis and healthy controls at p < 0.05.

|  | Healthy controls<br>(n=17) | MS patients<br>(n=28) | p-value | p-value<br>FDR<sub>correc</sub> |
| --- | --- | --- | --- | --- |
| **Demographics** |  |  |  |  |
| Sex, F/M<sup>•</sup> | 10/7 | 23/5 | 0.086 | 0.172 |
| Age | 39.29 (7.34) | 41.75 (10.01) | 0.385 | 0.420 |
| Disease duration |  | 6.50 (3.94) |  |  |
| Dominant hand, R/L<sup>•</sup> | 14/3 | 27/1 | 0.108 | 0.185 |
| **Disability scores** |  |  |  |  |
| EDSS<sup>a</sup> |  | 1.5 (1.0, 1.5) |  |  |
| Pyramidal FSS<sup>a</sup> |  | 1.0 (0.0, 1.0) |  |  |
| Cerebellar FSS<sup>a</sup> |  | 0.0 (0.0, 1.0) |  |  |
| **Volumetric data** |  |  |  |  |
| WMV<sup>b</sup>, % ICV | 33.48 (3.78) | 29.74 (3.70)* | **0.002\*** | **0.012\*** |
| CGMV<sup>b</sup>, % ICV | 31.55 (3.09) | 29.45 (2.30)* | **0.014\*** | **0.034\*** |
| DGMV<sup>b</sup>, % ICV | 5.94 (0.81) | 5.29 (0.64)* | **0.005\*** | **0.015\*** |
| Ventricles<sup>b</sup>, % ICV | 2.06 (1.17) | 2.65 (1.28)* | 0.130 | 0.195 |
| Lesion volume, log mm<sup>3</sup> |  | 3.10 (0.64) |  |  |
| Spinal cord area, mm<sup>2</sup> | 69.81 (6.82) | 70.71 (10.27) | 0.752 | 0.752 |
| **Task parameters** |  |  |  |  |
| Upper limb lag, ms | 184.71 (113.75) | 216.07 (90.08) | 0.201 | 0.268 |
| Upper limb force error, N | 0.31 (0.07) | 0.34 (0.10) | 0.296 | 0.355 |
| Lower limb lag, ms | 142.35 (116.49) | 266.43 (120.68)* | **0.002\*** | **0.008\*** |
| Lower limb force error, N | 0.30 (0.05) | 0.45 (0.16)* | **0.001\*** | **0.012\*** |

### Motor task performance

PwMS displayed a higher force error (+0.15 N, *p*=0.001) and a longer lag (+124 s, *p*=0.002) during the lower limb task but not during the upper limb task (Table 1, Figure 3). In both pwMS and HC, no correlations between force error and lag were found for either upper or lower limbs and between upper and lower limb force error. Upper limb lag was significantly associated with lower limb lag in HC only (p<0.001, Beta_std_=0.872) (Figure 3). No correlations were observed between task parameters and EDSS.

**Figure 3.**
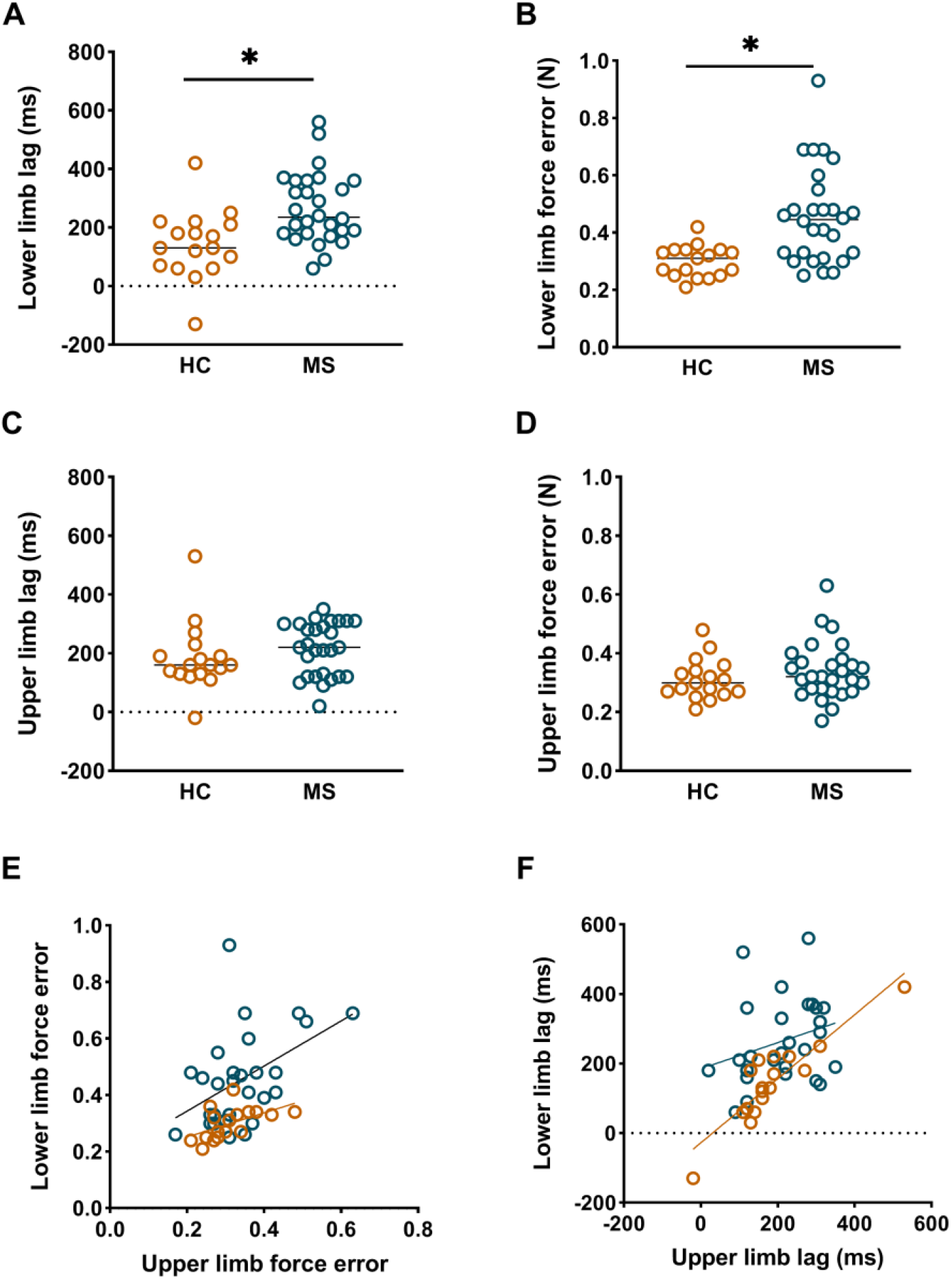
The functional force tracking task parameters. These plots demonstrate the group differences in task performance (lag and force error) and the correlations between the upper and lower limb performance. Each circle reflects a participant and the healthy controls (HC) are visualised in orange and people with multiple sclerosis (MS) in blue. **A-B)** People with MS showed significantly longer lag (+124 s, *p*=0.002) and higher force error (+0.15 N, *p*=0.001) during lower limb force tracking, compared to HC. **C-D)** No differences in performance were observed during upper limb movements. **E)** Force error of the upper and lower limb did not significantly correlate in both groups. **F)** Upper and lower limb lag correlated significantly in HC (*p*<0.001, Beta_std_=0.872), but not in MS (*p*=0.320, Beta_std_=0.210).

### Disease related functional activation changes

Main patterns of activation are shown in Figure 4. Both the upper and lower limb tasks resulted in large overlapping regions including the primary motor cortex (M1), premotor, supplementary motor area (SMA), primary somatosensory cortex, and secondary sensory cortex, cerebellar, deep grey matter (thalamus, putamen and pallidum). Besides sensorimotor regions, areas overlapped as well in the superior parietal cortex which is involved in visual spatial orientation.

**Figure 4.**
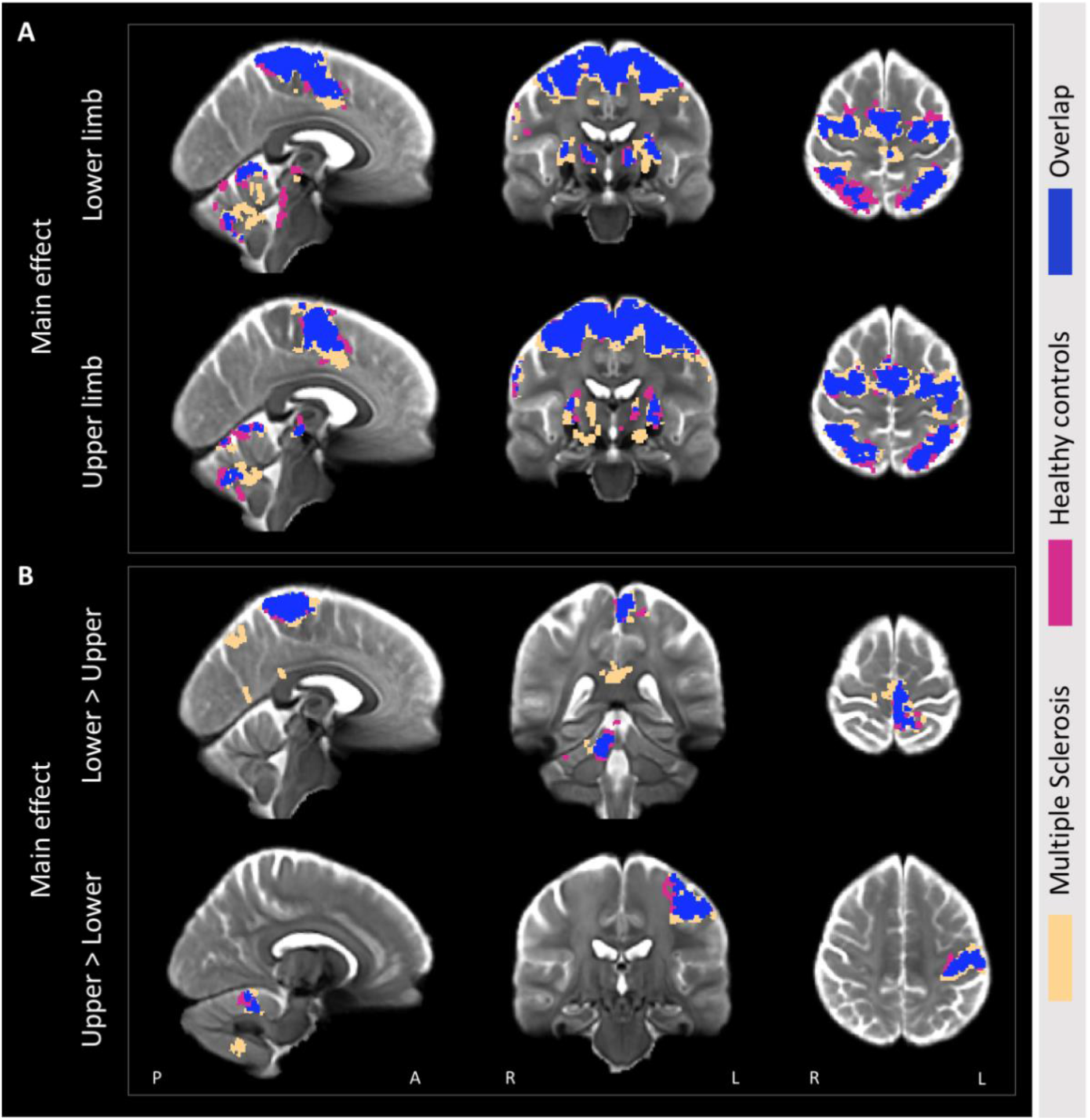
Average activation maps. **A)** The mean activation maps during the upper and lower limb visually guided force-matching tasks. **B)** Activation patterns specifically related to either lower limb (lower > upper) or upper limb (upper > lower) movements. The main effects for healthy controls (magenta) and people with multiple sclerosis (magnolia) are presented in an overlayed manner with the overlap between groups presented in dark blue.

During the lower limb task, pwMS displayed greater activation in contralateral (left) M1 and premotor cortex and reduced activation in the ipsilateral (right) cerebellar Crus I/II compared to HC (Figure 5). No activation changes were observed between groups during the upper limb task. No significant post hoc correlations were identified between peak z-stat values extracted from significant clusters and task performance variables, brain volumes, lesion volume or EDSS.

**Figure 5.**
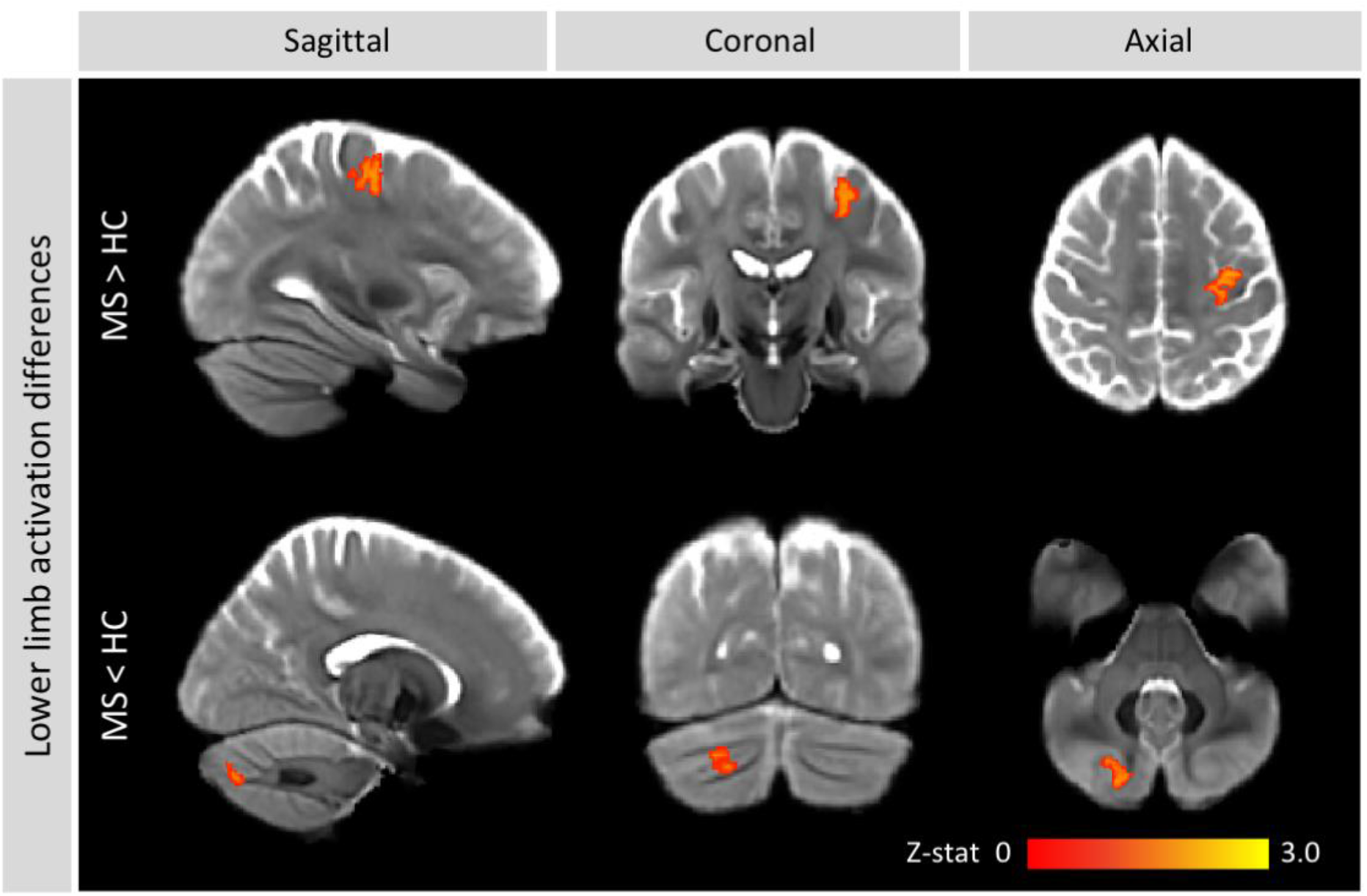
Lower limb functional activation changes in people with multiple sclerosis. During the lower limb force-matching task, people with multiple sclerosis displayed greater activation in the contralateral primary motor cortex and premotor cortex and lower activation within cerebellum Crus I/II, compared to healthy controls.

### Activation patterns related to upper and lower limb movements specifically

In both pwMS and HC activity specific to lower limb movements was observed in the contralateral medial primary somatosensory cortex and M1, whereas activation patterns related to upper limb movements were observed in lateral precentral gyrus, consistent with known somatotopy. Within the cerebellum, activity specific to upper limb movements was seen largely in the ipsilateral V-VI and lower limb activation was observed in the ipsilateral I-IV. Whereas the activation in primary sensorimotor cortices and the cerebellum largely overlapped between groups, pwMS recruited additional regions during both tasks compared to HC. In pwMS, activation specifically related to lower limb movements was also observed in the precuneus cortex, occipital cortex, frontal gyrus and cingulate gyrus. During the upper limb task, fewer additional regions were recruited, but in comparison to HC, pwMS recruited visual and cerebellar regions (Figure 4). These patterns of activation did not significantly differ between pwMS and controls.

### Task activation correlates of behavioural performance

Significant correlations between activation and task performance were found in several regions for the MS group, described in Figure 6 and Table 2. A positive correlation was displayed between lower limb task activation and lower limb lag in the contralateral SMA (medial Brodmann area 6), indicating that worse performance was related with higher activation. In addition, a negative correlation was seen in the contralateral parietal cortex (BA 5) and ipsilateral supramarginal gyrus (BA 40), indicating worse performance was correlated with lower activation. Force error correlated positively in the contralateral middle temporal visual area (MT). Upper limb task activation correlated negatively with upper limb force error, in the contralateral upper limb region of the M1 (BA 4), ipsilateral premotor cortex (dorsolateral BA 6) and left cerebellar VI.

**Figure 6.**
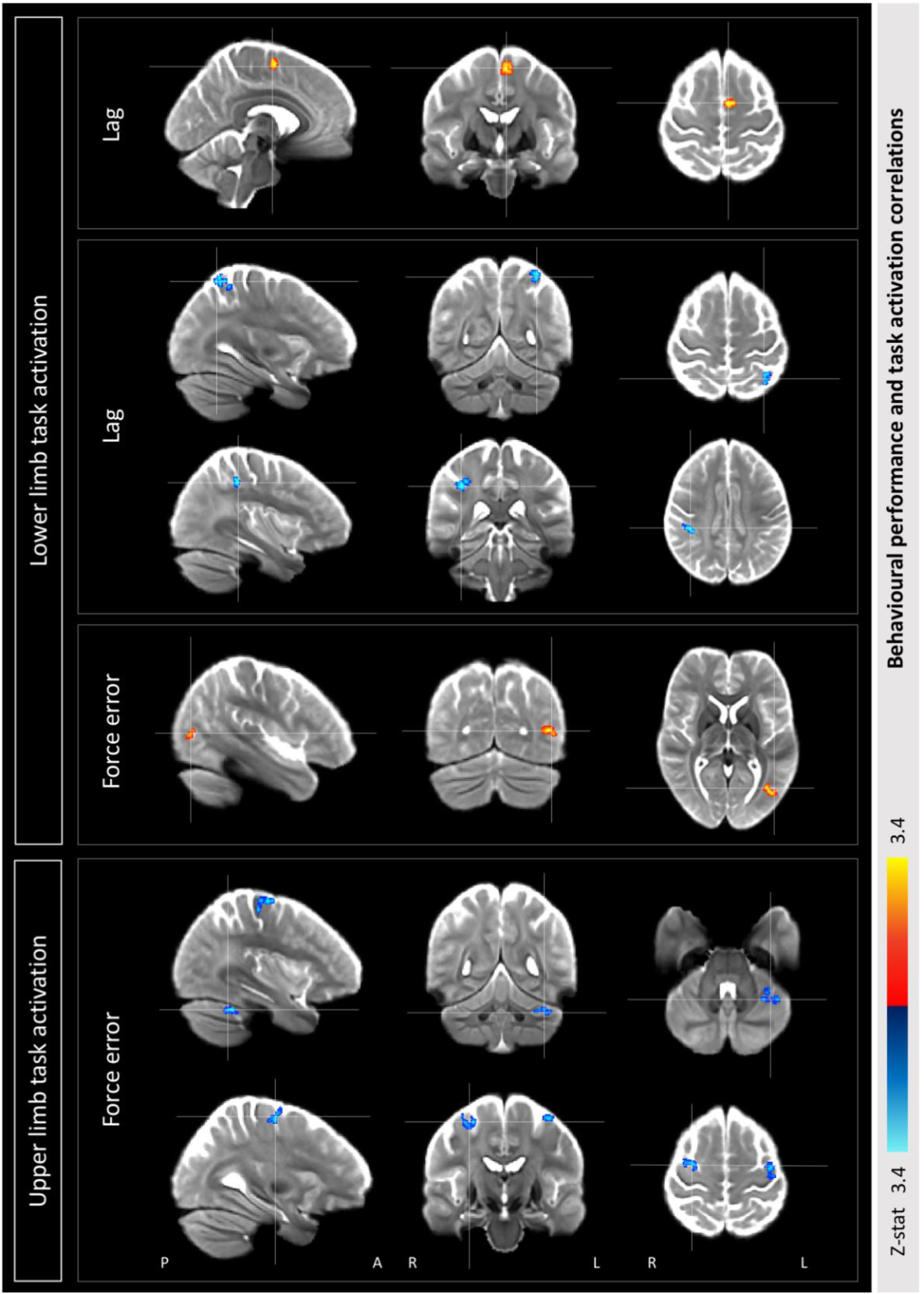
Task performance and activation correlations in multiple sclerosis. Positive (red) and negative (blue) correlations were identified between the behavioural measures lag and force error and task activation in cerebellar, visual and motor areas in people with multiple sclerosis.

**Table 2.**
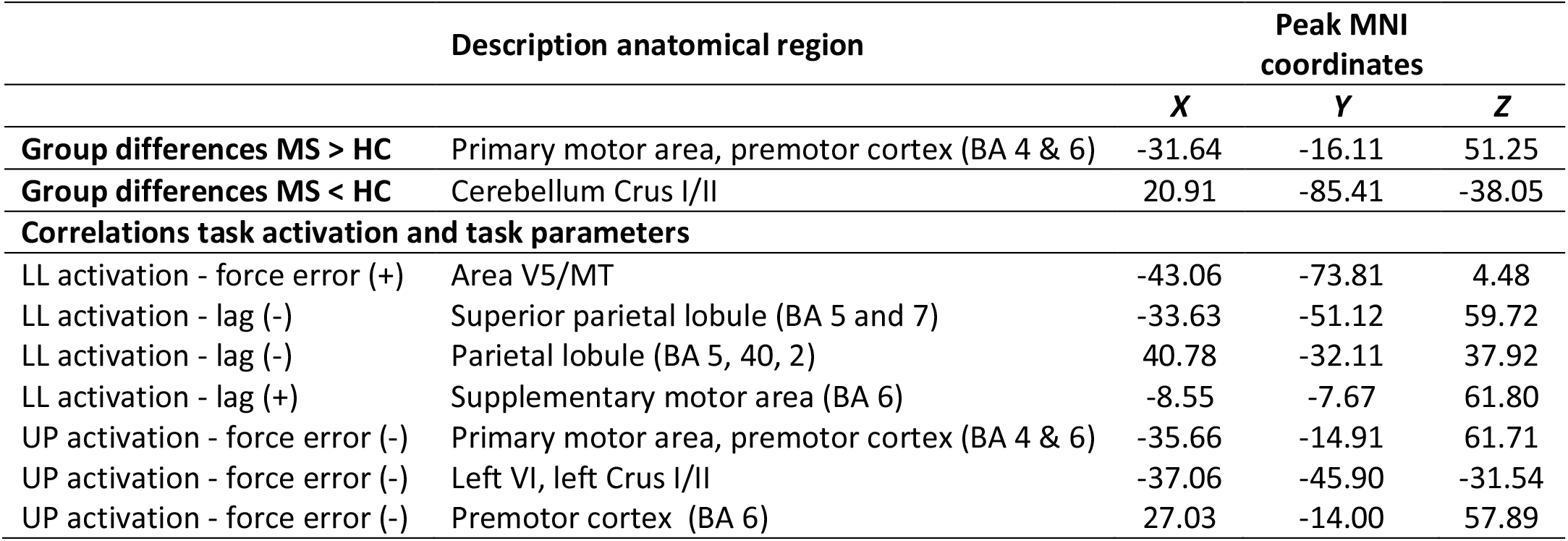
MNI coordinates and anatomical description. This table shows the location of the max z-stat of the significant clusters (group differences and correlations) in MNI coordinates. Cortical and subcortical regions are localised using the Brainnetome atlas(Fan et al., 2016) and cerebellar regions using SUIT (included in FSL 6.0.1, FMRIB 2012, Oxford, UK, https://fsl.fmrib.ox.ac.uk/fsl/). LL = Lower limb; UL = Upper limb; BA = Brodmann area.

## Discussion

In the present study, we aimed to investigate the complex upper and lower limb motor control in people with multiple sclerosis (pwMS) with minimal sensorimotor impairments using ultra-high field fMRI and a visually-guided force-matching task. We found that pwMS displayed altered task performance and hyperactivation of M1 and premotor cortex, but hypoactivation of cerebellum Crus I/II, during lower limb movements only, compared to healthy controls (HC). Upper limb task activation and motor performance was preserved and motor performance of the upper and lower limb did not correlate. Upper and lower limb task performance were correlated with activation in cerebellar, visual and motor areas.

### Lower limb functional activation changes

PwMS displayed increased cortical (M1, premotor cortex) and reduced cerebellar (Crus I/II) activations during lower but not upper limb force matching. These regions were adjacent to, but not within somatotopically mapped lower limb regions identified in the lower > upper contrast and therefore cannot be attributable to reduced sensory input or motor output to lumbar spinal cord motor neurons. Increased activation in M1 has been described in previous functional imaging studies in MS(Lee et al., 2000; Reddy et al., 2000, 2002; Rocca et al., 2005) and the primary sensorimotor cortices including M1 and S1 seem to be the first areas to show altered activation in the disease.(Filippi et al., 2004; Rocca et al., 2005) Increased premotor cortex activation has previously been reported, but either no significant between-group differences were found,(Lee et al., 2000; Reddy et al., 2000) or activation changes were observed ipsilaterally during hand movements.(Reddy et al., 2002) The increased activation could be a compensatory mechanism in response to the pathological damage, or perhaps a pathological loss of inhibitory control. However, to determine whether changes in activation are adaptive or maladaptive and are predictive of progressive decline would require longer term follow-up that was outside the scope of the current study.

In addition to cortical changes, we also observed reduced cerebellar Crus I/II activation during lower limb movements. In pwMS, reduced cerebellar activation has been observed during upper and lower limb motor tasks compared to controls.(Ciccarelli et al., 2006; Filippi et al., 2004; Mezzapesa et al., 2008; Pantano et al., 2005) A previous study reported contralateral reductions in the cerebellum in primary progressive MS patients with worse mobility,(Ciccarelli et al., 2006) whereas we observed ipsilateral changes in RRMS patients. Another lower limb study reported activation within the cerebellum but did not find significant differences between MS and HC,(Filippi et al., 2004) possibly due to subtle changes not measurable at the clinical field strength used. Cerebellar Crus I/II are suggested to be primarily involved in cognitive processes, as this region showed higher activation during a cognitive task compared to a finger tapping motor task(Stoodley, Valera, & Schmahmann, 2012), and have been included in cerebral networks involving cognitive control.(Buckner, Krienen, Castellanos, Diaz, & Yeo, 2011) However, lesions in these regions in stroke patients have been associated with worse motor task performance.(Stoodley, MacMore, Makris, Sherman, & Schmahmann, 2016) The link between Crus I and II and cerebrum association networks(Buckner et al., 2011; Stoodley & Schmahmann, 2010) and particularly the connection to the parietal lobe cortex(Stoodley & Schmahmann, 2010) together with the complex task used in this study suggests a potential role for these regions in dysfunctional visuomotor integration.

In contrast to our results, previous task fMRI studies have also reported altered activation during performance of upper limb tasks,(Lee et al., 2000; Mezzapesa et al., 2008; Pantano et al., 2002; Reddy et al., 2000; Rocca et al., 2005, 2003) but these studies included participants with longer disease durations, different MS phenotypes,(Lee et al., 2000; Reddy et al., 2000) or used a simpler motor task.(Lee et al., 2000; Mezzapesa et al., 2008; Pantano et al., 2002; Reddy et al., 2000; Rocca et al., 2005, 2003)

### Lower and upper limb performance in pwMS with minimal impairments

In our study, pwMS displayed delayed and more erroneous force tracking compared to HC for lower but not upper limbs. Also, we did not detect a significant correlation between tracking performance in upper and lower limbs in patients, despite observing such correlation in HC. This suggests that, at least in part, divergent pathological processes drive progression of upper and lower limb impairments. This interpretation is supported by a previous study showing only a moderate correlation between a hand-to-mouth task and walking revealed using advanced kinematic analyses.(Coghe et al., 2019) Also, clinical observations of pwMS often report impairments in lower limb function early in the disease.(Benedetti et al., 1999; Kister et al., 2013; Martin et al., 2006)

### Pathological factors influencing upper and lower limb functional impairments

The aim of this study was to investigate functional impairments in upper and lower limbs so we did not undertake detailed structural investigations. However, we did identify a gross atrophy of cortical and deep grey matter and white matter. Possibly structural damage within the cerebrum might explain the activation and behaviour changes observed during lower limb movements specifically. The thalamus is one of the first regions to become atrophic followed by the precentral gyrus and cerebellum in relapse-onset disease,(Eshaghi et al., 2018) possibly leading to activation changes in M1. Interestingly, we did not observe differences in cervical spinal cord cross-sectional area between patients and controls. We were unable to fully characterise cervical cord lesions due to the limited field of view afforded by the use of a transmit/receive head coil necessary for 7T imaging. However, lesions are primarily observed in the cervical spinal cord (59%),(Weier et al., 2012), clearly affecting both upper and lower limb function and early MS spinal cord lesions are often asymptomatic(Granella et al., 2019) and do not predict disability progression(Dekker et al., 2017).

### Force-matching for studying visuo-motor integration in pwMS

Here we used a force matching task to elicit submaximal contractions of the tibialis anterior (lower limb task) or hand flexion (upper limb task) performed in separate runs adapted from a task used previously to investigate motor cortex reorganisation in knee osteoarthris.(Shanahan et al., 2015) With this task we aimed to model not only basic sensorimotor behaviour, but complex integrative sensorimotor behaviour required for daily life. Similar tasks have been used previously to study the basic neurophysiology involved in visuomotor control.(Keisker, Hepp-Reymond, Blickenstorfer, & Kollias, 2010; Mayhew et al., 2017) These studies showed that visual feedback similar to that used here resulted in stronger activation, and that activation scaled with the MVC. However, stronger correlations were found between activation and behaviour performance when participants performed at 10% MVC compared to 30%, potentially due to greater neuronal recruitment required for finer movement control at lower MVC. We used a MVC of 5% which was selected to both increase the difficulty of the task, and to minimise the risk of head motion during tibialis anterior contractions. While apparently straightforward, the task activated a broad network of visual attention and oculomotor areas as well as visuomotor integration, premotor and primary sensorimotor cortices. The patterns of activation were almost identical for upper and lower limb tasks, with only somatotopically mapped contralateral M1/S1 and ipsilateral cerebellum differentiating the two tasks. We conclude that force-matching tasks provide a relatively straightforward means to investigate complex visuomotor integration with functional MRI.

### Methodological considerations

In this study we used ultra-high field MRI with the goal of detecting more specific activation loci with better accuracy due to high sensitivity. For low resolution fMRI data, large spatial smoothing kernels are commonly used to enhance SNR and to minimise the effects of misregistration. However, this also reduces accuracy as it can lead to incorrect estimation of true spatial localisation and therefore type-I error.(Heidemann et al., 2012; Sacchet & Knutson, 2013) We therefore chose to create a study-specific template to improve the accuracy of spatial coregistrations and therefore we were able to use very limited smoothing in our data (2.5 mm). To conclude, using ultra high field and a study-specific template led to identification of detailed activation patterns specific to movement without losing sensitivity.

Even though subjects only performed the task with the right hand and foot, 3 of 45 subjects (1 MS, 2 HC) were left handed. Given the small proportion of left handers, we do not expect dominance to influence results significantly. While there are significant advantages for the use of ultra-high field for functional MRI such as higher raw signal to noise ratio(Yoo et al., 2018), stronger susceptibility effects(Ladd et al., 2018), and signal more closely attributable to venules than feeding veins(Cheng, 2018)), worse magnetic field inhomogeneities can lead to signal dropout and warping around the inferior temporal and orbitofrontal cortices. For the current study these areas were not of major interest and the study-specific template demonstrated the very high SNR across the brain with very little signal loss in areas of interest such as the sensorimotor cortices and cerebellum.

## Conclusion

These results demonstrate that ultra-high field fMRI during complex hand and foot tracking can identify subtle impairments in movement and brain activity in otherwise minimally impaired pwMS, with differential effects for upper and lower limb impairments. Minimally disabled pwMS displayed altered lower limb movements and brain activation with preserved upper limb function and activation. Such techniques might find use in tracking and predicting the emergence of impairments in mobility and dexterity that reflect some of the most disabling symptoms of MS.

## Data Availability

Anonymised data is available from the corresponding author upon request.

